# Wastewater-integrated pathogen surveillance dashboards enable real-time, transparent, and interpretable public health risk assessment and dissemination

**DOI:** 10.1101/2025.01.22.25320865

**Authors:** Nosihle S. Msomi, Joshua I. Levy, Nathaniel L. Matteson, Nkosenhle Ndlovu, Phindile Ntuli, Adam Baer, Victor Mabasa, Sipho Gwala, Natasha Singh, Kathleen Subramoney, Emmanuel Phalane, Mokgaetji Macheke, Mantshali Motloung, Thabo Mangena, Lethabo Monametsi, Lebohang Rabotapi, Sibonginkosi Maposa, Amanda Birmingham, Mark Zeller, Smruthi Karthikeyan, Simon Harris, Rob Knight, Louise C. Laurent, Kristian G. Andersen, Kerrigan McCarthy, Mukhlid Yousif

## Abstract

Timely pathogen surveillance and reporting is essential for effective public health guidance. Web dashboards have become a key tool for communicating public health information to stakeholders, health care workers, and the broader community. Over the SARS-CoV-2 pandemic, wastewater surveillance has increasingly been incorporated into public health workflows for outbreak monitoring and response, enabling community-representative and low-cost monitoring to supplement clinical surveillance. However, the methods used for visualization and dissemination of clinical and wastewater surveillance data differ across programs and best practices are yet to be defined. In this work, we demonstrate data workflows and dashboards used to perform wastewater-based public health surveillance in tandem with clinical data across local and national scales, leveraging custom-built, reproducible, and open-source software. Using a centralized data aggregation and analysis hub approach, we establish multiple data pipelines for data storage, wrangling, and standardized analyses, and deploy custom-built web dashboards that allow for immediate public release. We find that our approach is effective across scales, computing architectures, and dissemination strategies, and provides an adaptable model to incorporate additional pathogens and epidemiological data.

## 1. Introduction

In recent years, wastewater-based pathogen surveillance has been widely adopted as a supplement to clinical case-based surveillance efforts (1–3). Wastewater surveillance is cost-effective, limits sampling biases, and can be used to identify outbreaks, quantify pathogen levels, and track circulating variants (4,5). However, sharing of wastewater surveillance data and derived insights with the public presents additional technical demands and dissemination considerations for already resource-limited public health organizations(6,7). To effectively communicate complex and often multimodal epidemiological findings in a timely manner, many public health organizations have begun to deploy real-time, interactive digital dashboards (8).

Pathogen surveillance dashboards visually synthesize data into key metrics and graphs to provide streamlined communication with public health stakeholders and members of the public (8–10). Open data access via a dashboard allows for transparent public health guidance and intervention and empowers individuals to make educated decisions about their health and behaviour (11–13). In practice, maintaining a real-time dashboard requires close coordination of sample collection, laboratory processing, and computational efforts to analyze and visualize data and derived metrics, including clinical cases and deaths (14), epidemiological projections (15), wastewater pathogen prevalence and lineage dynamics (16).

In this study, we describe the steps needed to build integrated clinical and wastewater surveillance dashboards and the core data architectures and workflows used for dashboard development and deployment. We deploy these approaches in support of wastewater-integrated SARS-CoV-2 surveillance in South Africa and in San Diego County, USA, allowing for public communication of key findings including wastewater viral loads, variant prevalence, and reported clinical cases in real-time. We establish core design principles for dashboard development and structure and provide reusable dashboard templates that are freely available for community use.

## 2. Methods

### Data collection and aggregation

Wastewater grab samples are collected from 50 wastewater treatment plants (WWTPs) and sub-catchment areas across South Africa each week. Sample collectors use the RedCap Mobile App (17) to capture sample metadata during sample collection. Samples are then sent to the Centre for Vaccines and Immunology at the NICD, where they are tested for SARS-CoV-2 using a digital PCR (dPCR) assay. The dPCR results are also captured on RedCap. Samples tested positive are sequenced using a whole genome amplicon sequencing approach (5).

Since COVID-19 is classified as a notifiable medical condition in South Africa, private and public laboratories throughout the country are required to report any suspected and laboratory confirmed cases. These clinical case data are stored in the National Institute for Communicable Diseases’ (NICD) Surveillance Data Warehouse (SDW) (18). For the purpose of the dashboard, only dPCR laboratory confirmed cases from the public and private health sector in districts where wastewater samples are collected are analyzed. Accompanying clinical case counts are included alongside wastewater data.

Custom R scripts are used to anonymize all clinical data, perform data wrangling of both the clinical and wastewater data and transform the data into a format that can be easily used for the dashboard. The data are then placed in a publicly accessible GitHub repository which may be accessed online (https://github.com/NICD-Wastewater-Genomics/NICD-Dash-Data). The R script uses an API to connect to RedCap, and connects to the SDW to get up-to-date information each time the script is run. This script is scheduled to run daily within a linux server using a cron job scheduler.

The San Diego Epidemiology and Research for COVID Health (SEARCH) alliance collects wastewater from three regional WWTPs, Point Loma, South Bay, and Encina using a composite sampling method. Viral loads are estimated using RT-qPCR and normalized by pepper mild mottle virus (PMMoV) concentration, and whole genome amplicon sequencing is performed on all positive samples. From February 2021 to May 2023, accompanying clinical case counts were obtained via a public data stream provided by San Diego County and included alongside wastewater data (https://www.sandiegocounty.gov/content/sdc/hhsa/programs/phs/community_epidemiology/dc/2 019-nCoV/status/COVID19_Cases_by_Geography_of_Residence.html). Sewer maps, available through the San Diego County Open Data Portal (https://gis-sangis1.hub.arcgis.com/pages/download-data) are used to determine the zip codes contributing to each plant. Zip code-level case counts are also available from the Data Portal and are summed by catchment.

Viral load estimates are directly uploaded to a public GitHub repository (https://github.com/andersen-lab/SARS-CoV-2_WasteWater_San-Diego) by the UCSD EXCITE laboratory. Lineage prevalence estimates from sequencing analyses are uploaded to the same repository following bioinformatic analysis on Scripps Research and UCSD computing resources. Basic unit testing and quality control tests, followed by manual assessment of results, are used to check for possible issues with data uploads prior to inclusion in public data visualizations.

### Computational and bioinformatic workflow

SARS-CoV-2 sequences generated from wastewater samples are analyzed using Freyja as previously described (5). Freyja output files produced during sequence analysis are also placed in a publicly accessible GitHub repository (https://github.com/NICD-Wastewater-Genomics/NICD-Freyja-outputs-). Automated workflows using GitHub Actions are used to update outputs in line with available lineage designations. All dashboard data files are aggregated to a single centralized dashboard data repository (https://github.com/NICD-Wastewater-Genomics/NICD-Dash-Data), using automated cronjobs. The SEARCH dashboard and the NICD v2 dashboards are built using a custom Plotly Dash script(19), while the NICD v1 dashboard was built using Tableau. To improve page download and loading times, all data for the NICD dashboard is compressed into feather format, a more compressed, fast-loading binary storage format for matrices and data frames (20). For each data update, growth rate calculations using a logistic model are performed automatically using a GitHub action workflow.

### Normalization, binning, and smoothing

In South Africa, nationwide viral load estimates are calculated by averaging viral loads from all sites in South Africa for each epidemiological week (epiweek). Clinical data are similarly summed by epiweek. Viral load curves are smoothed using a linear, non-uniform sampling Savitzky-Golay filter, while lineage prevalence trends are smoothed using a simple moving average filter. In San Diego, all analyses are focused on a single catchment, and smoothing of viral loads, clinical curves, and lineage prevalences are done using a linear Savitzky-Golay filter.

### Serving an online dashboard

The SEARCH dashboard is served using independent HTML iframes, directly incorporated into the SEARCH website, which is designed using Wordpress. The Dash application is deployed using Amazon Web Services (AWS) Elastic Beanstalk, using a t2.small instance and embedded into the SEARCH website using an iframe. The NICD v2 dashboard is served as a standalone, multi-page website using an in-house web server configured with a virtual machine running Ubuntu 24.04, at least 4 vCPUs, 8GB of RAM, and 150GB of storage.

### Ethics declarations

The wastewater surveillance work in San Diego was discussed with the University of California San Diego Institutional Review Board, and was not deemed to be human subject research as it did not record personally identifiable information. All clinical case counts for San Diego were obtained via openly available, anonymized data streams provided by the County of San Diego. The protocol for the work performed in South Africa was reviewed and approved by the University of the Witwatersrand Human Research Ethics Committee (MM220904). The NICD conducts all routine clinical surveillance, including surveillance of notifiable medical conditions, in a protocol reviewed and approved by the University of the Witwatersrand Human Research Ethics Committee (HREC) M210752 and under the legal authority of the National Health Act (no. 61 of 2003).

## 3. Results

### Maintaining data and processing infrastructure for dashboards

To facilitate interpretable and robust data analyses, we use multi-stage, distributed data processing and updating workflows (Figure 1). For both the SEARCH and NICD programs, separate workflows for wastewater and clinical sample collection, analysis, and reporting are integrated into a unified visualization. Wastewater samples are processed in a single lab to obtain viral load measurements and perform sequencing. Subsequently, the data are uploaded to a public data repository, where samples can be automatically re-analyzed and outputs updated following designation of new virus lineages. In parallel, clinical case counts obtained in collaboration with diverse networks of labs, practitioners, and public health agencies.

**Figure 1:**
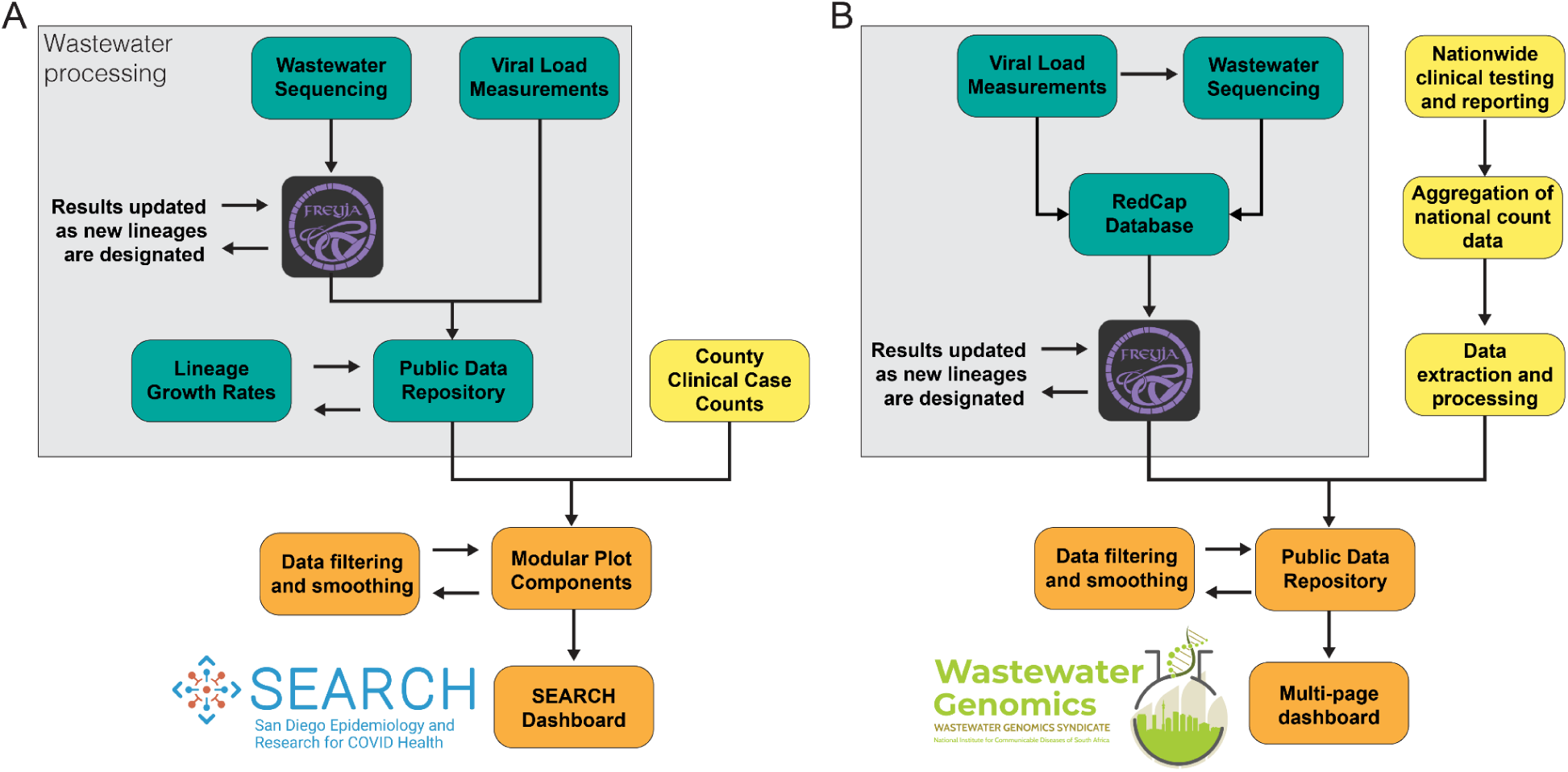
Distributed data analysis and multi-source integration. A. For the SEARCH dashboard, separate analysis pipelines are used for wastewater sequencing data and viral load measurements, but both are uploaded to a public data repository, triggering automated recalculation of lineage growth rates. Clinical case data are pulled from county data resources,integrated within plotting modules, and inserted individually into the SEARCH website. B. For the NICD dashboard, viral loads in wastewater samples are first quantified to determine if they can be sequenced. The results are added to a RedCap database. Sample sequences are analyzed with Freyja and uploaded to a public data repository alongside clinical case counts obtained through the NICD’s surveillance data warehouse. Data from the public data repository is pulled by the dashboard when loaded by the user.

Wastewater and clinical data streams are matched for each region and filtered or smoothed in time to highlight trends present in the raw data. For sequencing analyses, lineage prevalence estimates are summarized into custom groupings of leading virus variants and their sublineages, including both locally identified variants as well as CDC and WHO designated variants. For transparency, all wastewater measurements and results are stored on public GitHub repositories, where they are time-stamped and version controlled.

### SEARCH Wastewater Dashboard

SEARCH began wastewater surveillance in July 2020 in response to a need for a secondary mechanism for tracking of COVID-19 burden across San Diego County (21,22). The wastewater program is run in parallel with SEARCH clinical surveillance (23), and covers three WWTP serving over 2.6 million individuals, including nearly all metropolitan San Diego residents and approximately 80% of all county residents (Figure 2A).

**Figure 2:**
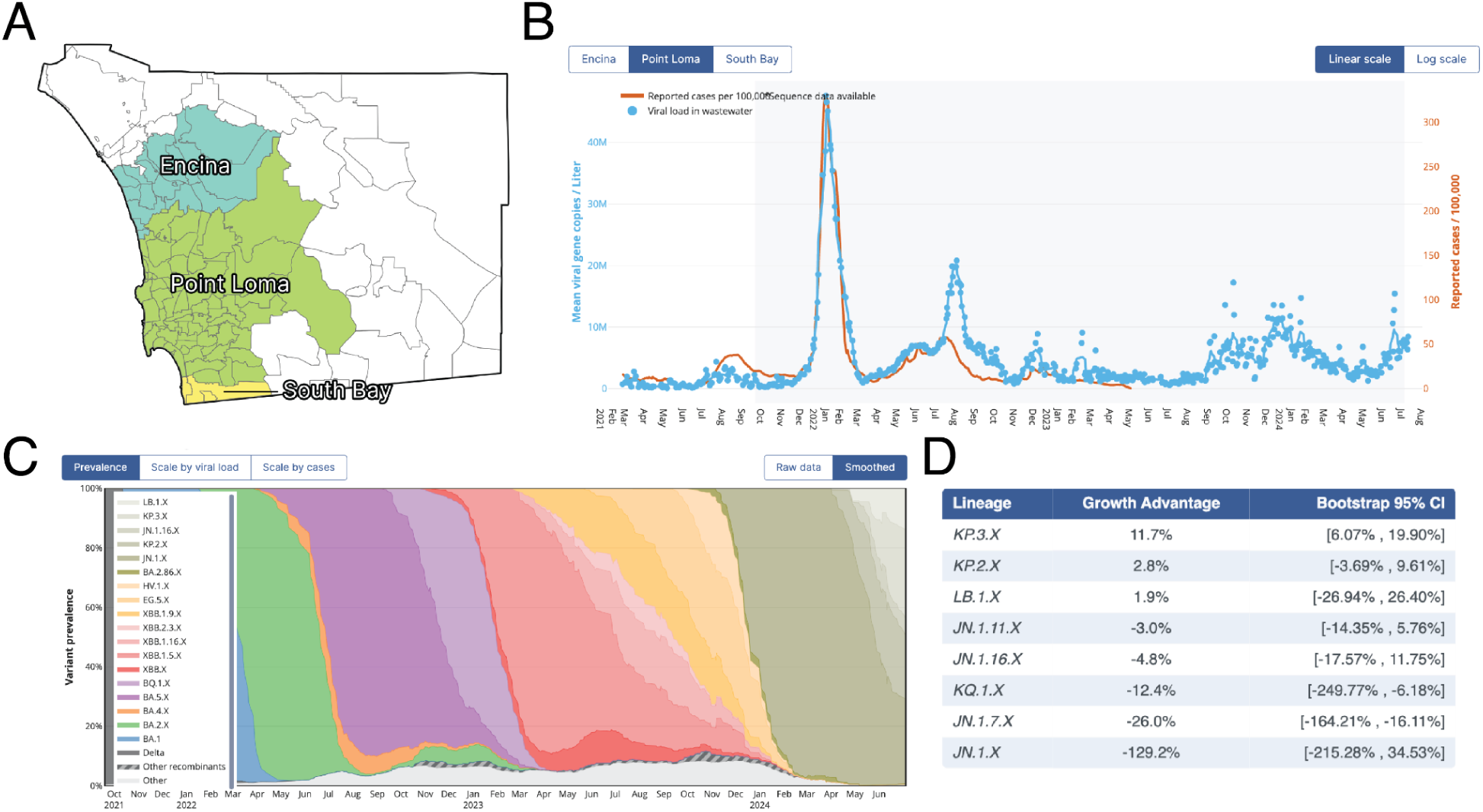
SEARCH Wastewater Dashboard components. A. Zip code-level mapping of catchment areas for each San Diego wastewater treatment plant. B. Combined visualization of wastewater viral loads and catchment specific-case counts. C. Lineage prevalence tracking for each catchment, with sub-lineages grouped with parent lineage. D. Relative growth advantage estimates for each lineage grouping, using circulating lineages as background.

The SEARCH dashboard (https://searchcovid.info/dashboards/wastewater-surveillance/) enables direct comparison of wastewater viral loads and clinical case counts, with the raw estimated viral loads measurements shown for user-interpretability (Figure 2B). Using a multi-button interface, users can navigate between each of the catchments and toggle the scaling of the visualization (Figure 2B). For sequencing data, longitudinal lineage prevalence tracking is presented using stacked area charts (Figure 2C), along with corresponding estimates of lineage growth rates to identify variants with outbreak potential (Figure 2D). Similar to the viral load trajectory component, there are interactive switches to reveal the raw data, or to scale the lineage prevalence estimates by viral load or case count, which provides an estimate of the total burden associated with each lineage grouping. The dashboard plots are modular and easily reused, which can be leveraged to rapidly deploy similar visualizations, including for other pathogens of public health concern. For example, during the 2022 multi-country mpox outbreak, tracking of clinical cases and wastewater viral loads was made available directly via a separate SEARCH dashboard (S1 Figure).

### NICD Wastewater Surveillance Dashboard

The NICD in South Africa adopted a SARS-CoV-2 wastewater surveillance programme in 2021 (4). This programme is run in parallel to clinical surveillance, acting as an early warning signal to changes in SARS-CoV-2 disease dynamics. As part of this surveillance effort, an initial public dashboard (NICD v1 dashboard) was developed by an external provider to display viral load trends observed at WWTPs across the country (S2 Figure).

To integrate clinical data, wastewater data and sequencing information, we developed an improved, simple, and easy-to-use custom dashboard. The resulting dashboard contains dedicated pages that provide an overview across the country as well as at a district level (https://wastewater.nicd.ac.za/). At the national level, data cards provide information about the current epidemiological week, the number of laboratory-confirmed SARS-CoV-2 clinical cases reported, and how many wastewater samples were collected (Figure 3A). SARS-CoV-2 viral load trends over time, along with a smoothed interpolant, are plotted alongside clinical case counts(Figure 3B). Users hovering on this graph are able to view the exact clinical case count and viral load for a given epiweek. A stacked area chart (Figure 3C) is used to visualize the prevalence of various SARS-CoV-2 lineages over time, and interactive switches allow users to switch between daily and monthly time resolution.

**Figure 3:**
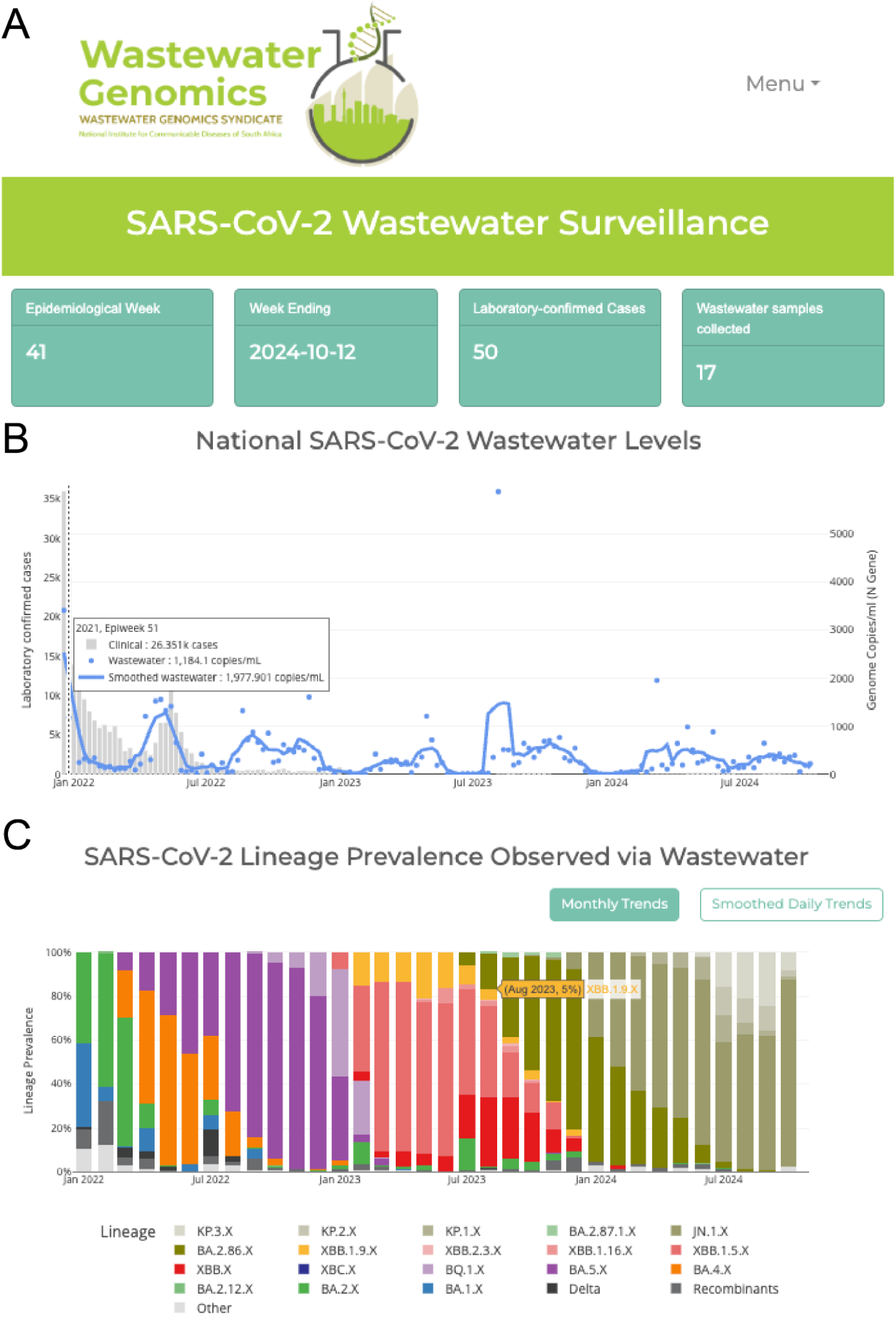
NICD National Wastewater Dashboard core components. A. Data cards provide information on the current epidemiological week as well as the clinical case counts and wastewater samples collected. B. SARS-CoV-2 viral load compared to the number of clinical cases reported each week, with interactive hover to see exact values in the chart. C. Lineage prevalence over time, with toggle option to switch between smoothed and binned data.

The NICD dashboard includes a secondary page for investigation of wastewater and clinical trends at the district level. Users can select a district of interest, which is interactively shown on a map of monitored sites across the country (Figure 4A). Alongside, the selected districts’ clinical cases are shown with wastewater viral load trends from each WWTP in the district (Figure 4B). When users hover above a point in the graph, they are able to view the date the sample was collected, the WWTP from which it was collected, and the viral load detected in that sample. Similarly to the national page, users can view the lineages detected at each WWTP as well as sub-catchment areas in Gauteng (Figure 4C).

**Figure 4:**
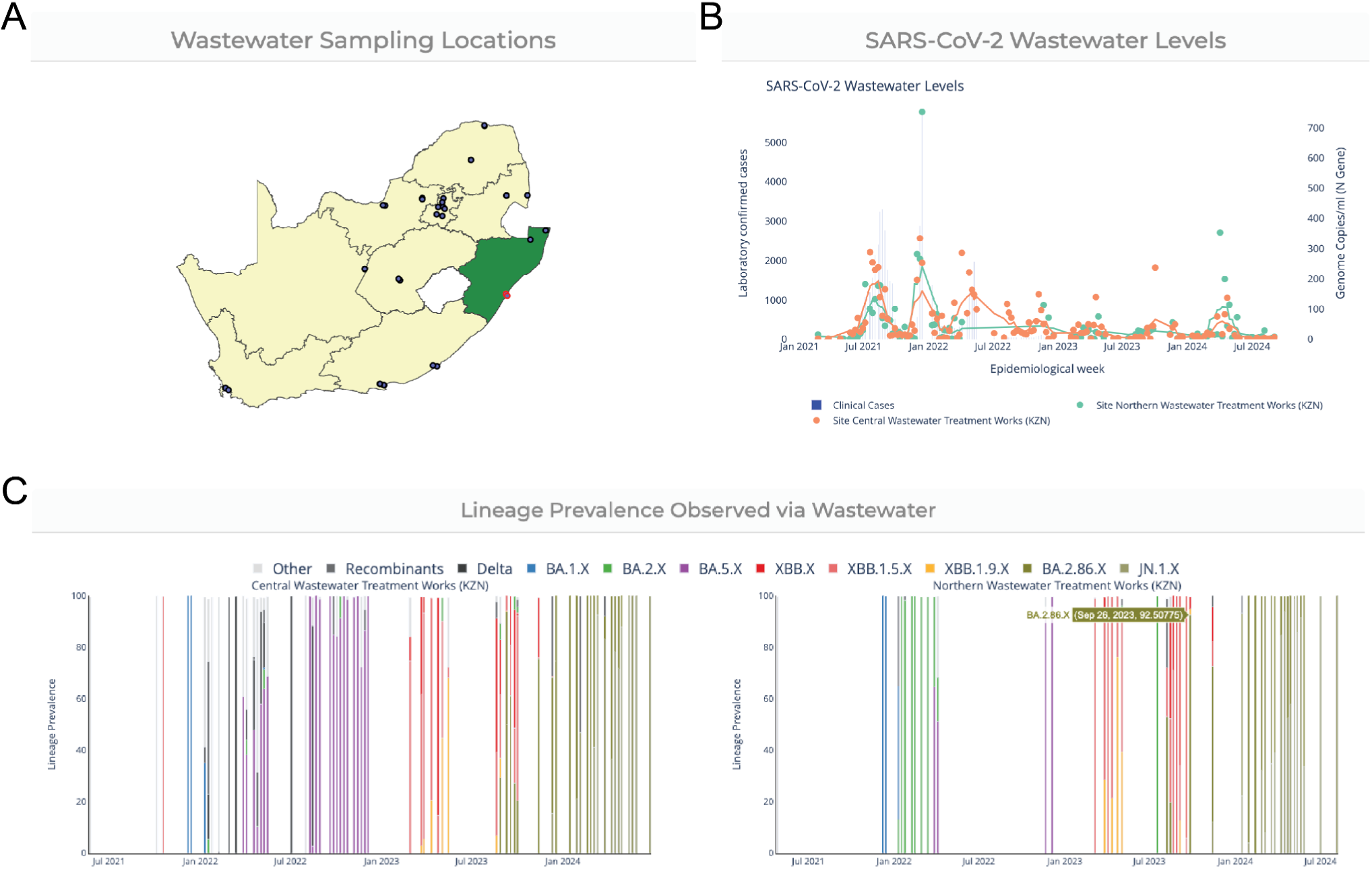
NICD Wastewater Dashboard district-level Components. A. Users can select a district of interest (highlighted in green), and the corresponding WWTPs within that district are highlighted in red. B. The viral loads for each WWTP in the district, along with district-level case counts, are shown over time. C. Lineage prevalence tracking for each WWTP, resolved by date of sampling.

## Discussion

Wastewater surveillance systems can provide real-time, community-representative information on pathogen evolution and spread (1). We present the NICD and SEARCH data workflows and dashboards to provide clean, modular, and extensible solutions for public health monitoring in support of pathogen surveillance from the city to national-scale. These tools enable rapid adoption of wastewater surveillance to supplement current clinical surveillance efforts, especially important in regions with limited access to clinical testing, where wastewater can identify and fill clinical surveillance gaps and guide response efforts(4,24).

Maintaining real-time dashboards can be technically demanding, generally requiring coordinated data collection, analysis, and visualization, as well as significant computational resources and expertise. Both NICD and SEARCH use open-source tools and platforms, such as R for data processing and GitHub for data storage and to coordinate data flows with limited need for expensive computing resources. However, these platforms need to be carefully managed to ensure that data are accurate, secure, and appropriately anonymized. The ongoing development of best practices and standards for data collection, storage, analysis, and dissemination, including by groups such as PHA4GE (25), WHO (26), and the Global Consortium for Wastewater and Environmental Surveillance for Public Health (27), is essential to ensuring data interpretability, accessibility, and protection of community privacy. Given that these data are inherently tied to specific communities, collaboration with local municipalities is essential. Beyond providing data through the dashboard, it is essential to engage with local stakeholders, meeting regularly and sharing detailed reports of recent findings to ensure that communities benefit directly from the sampling efforts.

Depending on contextual factors such as expected traffic, intended usage, and technical resources, the deployment platform and underlying software can vary significantly. For example, the SEARCH dashboard was designed to accommodate a potentially large and geographically dispersed audience, including researchers and public health stakeholders accessing the platform remotely. Using commercial cloud computing resources, SEARCH researchers directly managed scalable deployments without requiring specialized IT support. In contrast, in-house web server infrastructure was used to host the NICD v2 dashboard, aligning with organizational policies on data security and governance. This approach required technically skilled IT personnel for initial setup and maintenance, as well as effective coordination across institutional departments.

For users to effectively utilize and interpret the data aggregated on a dashboard, the dashboard needs to be easy to understand, appealing, reliable, and trustworthy (8). As such, both dashboards make use of interactive components such as toggles for the lineage prevalence and smoothing options for the viral load in order to enhance user engagement and understanding. Additionally, it must be clear that data provided by dashboards is timely and accurate. As soon as data are available, pre-validated processing workflows are automatically triggered to provide dashboard updates. Both NICD and SEARCH make use of live data feeds and automated workflows to ensure that users can access the most up to date information as soon as possible, and clearly indicate the time of the last update.

The sharing of customizable, reusable, low barrier of entry computational workflows and dashboards with the public health community can enable other labs to build similar dashboards for their communities and help establish core standards of practice. For this purpose, we provide open access to the source code for our complete workflows and dashboard architectures, including automated scripts and public repositories used for accurate and timely data integration. This codebase is sufficient to build scalable end-to-end workflows and can be modified to incorporate custom analyses or expansion to multiple pathogens as needed.

### Conclusion

Our approach for real-time and interactive dashboards for wastewater-integrated surveillance represents a key resource to increase robustness and expand access to public health monitoring. By integrating wastewater and clinical data, we show that dashboards can offer a comprehensive view of pathogen dynamics to support timely and informed public health decision-making. We establish robust and modular data workflows and visualizations for multi-modal pathogen surveillance and make them openly available to the broader research and public health community.

## Code availability

For the NICD dashboard, backend wastewater analysis code (https://github.com/NICD-Wastewater-Genomics/NICD-Freyja-outputs-) and custom R (v 4.3.3)(28) scripts used for sample anonymization are available via github (https://github.com/NICD-Wastewater-Genomics/In-house-scriipts/tree/main/R_script_for_Dashboard). The NICD web dashboard is written in python using the Plotly Dash package (v2.14.0) and is available through a separate repository (https://github.com/NICD-Wastewater-Genomics/Wastewater-Dashboard). For the SEARCH dashboard, processing scripts (https://github.com/andersen-lab/sd_ww_processing) and the dashboard website (https://github.com/andersen-lab/lone_pine) are all available in dedicated repositories.

## Data availability

All raw data used in this study is available through BioProjects PRJNA819090 and PRJNA941107, and as processed Freyja outputs in public repositories (NICD: https://github.com/NICD-Wastewater-Genomics/NICD-Freyja-outputs- and SEARCH: https://github.com/andersen-lab/sd_ww_processing).

## Acknowledgements

We would like to thank L. Asato and the Microbiology Laboratory at the SD Public Utilities Department for providing us with county wastewater samples, the SEARCH (San Diego Epidemiology and Research for COVID Health) Alliance, local government and wastewater treatment staff in South Africa for sample collection and transport, the staff of the NICD Centre for Vaccines and Immunology and the Centre for Respiratory Disease and Meningitis, and the members of the Modjadji wastewater surveillance initiative.

## Author Contributions

Conceptualization: NSM, JIL; Data Curation: NSM, JIL, NLM, NN, PN, AB, VM, SG, NS, KS, EP, MM, MaM, TM, LM, LR, AB, MZ, SK, KGA; Formal Analysis: NSM, JIL, NLM; Investigation: NSM, JIL, NLM, NN, PN, AB, VM, SG, NS,AB, MZ, SK; Methodology: NSM, JIL, NLM, NN, PN, AB, VM, SG, NS,AB, MZ, SK; Visualization: NSM, JIL, NLM; Software: NSM, JIL, NLM, NN, PN, AB, VM, SG, NS,AB, MZ; Funding Acquisition: RK, LCL, KGA, MY, KM; Resources: SH, SM, RK, LCL, KGA, MY, KM; Supervision: SM, RK, LCL, KGA, MY, KM; Project Administration: SM; Writing – Original Draft: NSM, JIL; Writing – Review & Editing: all authors.

## Supporting Information

**S1 Figure:**
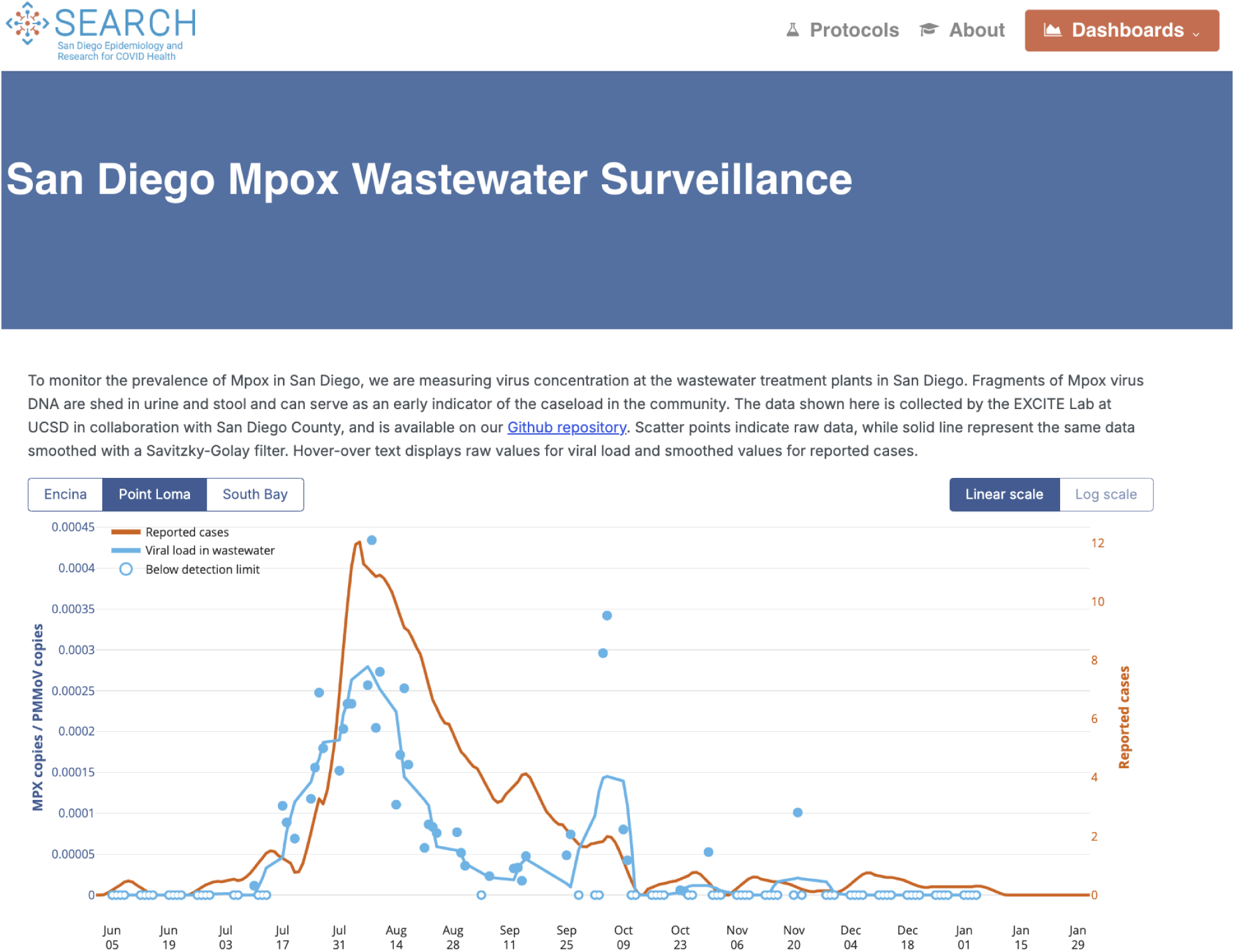
Mpox combined wastewater and clinical dashboard. Matched clinical and wastewater surveillance trends are provided for each of the three San Diego WWTPs, using the same approach used for the initial wastewater dashboard. This code is modular and resulting plots can be easily inserted as an iframe.

**S2 Figure:**
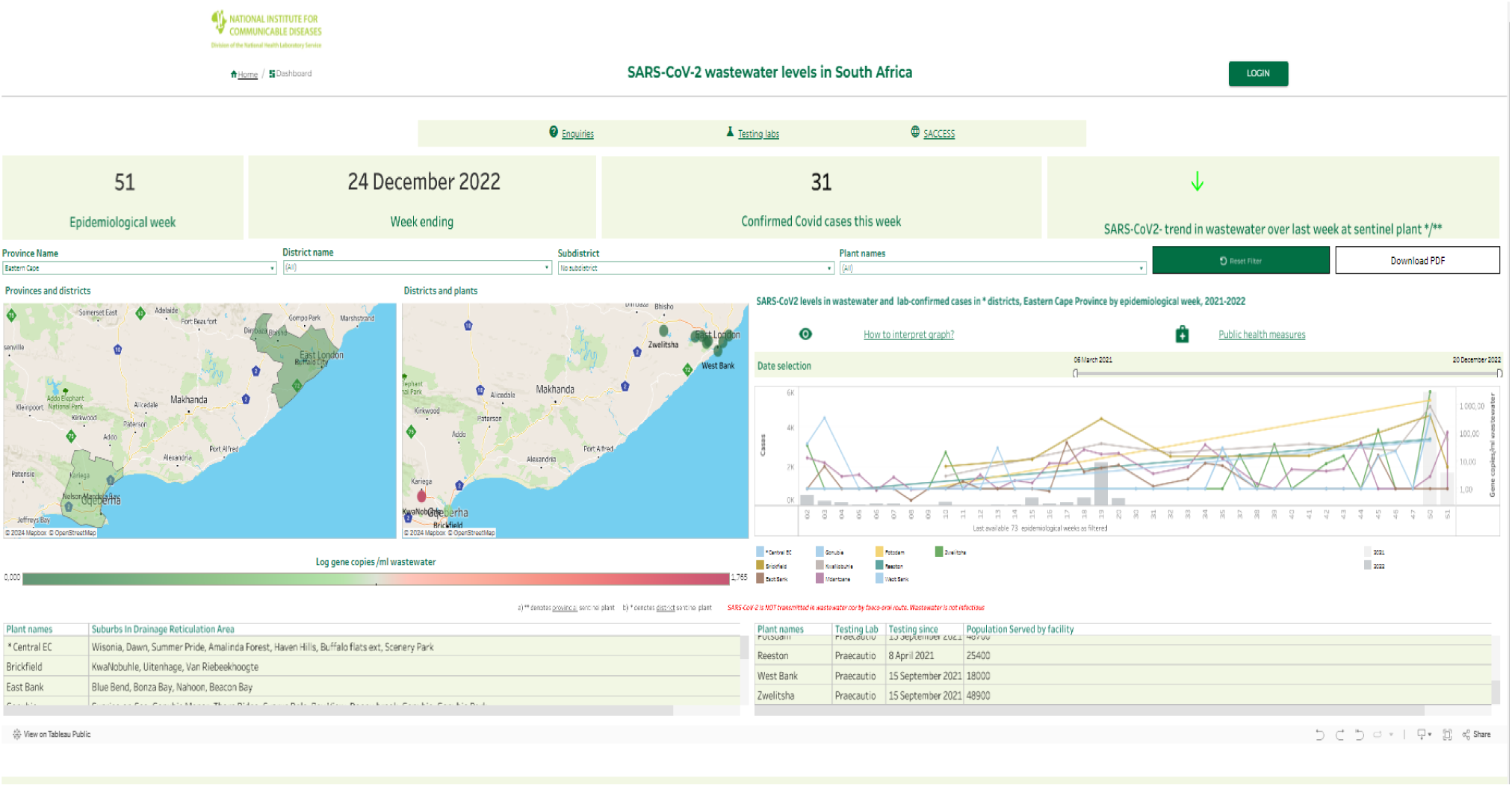
Initial NICD Dashboard. Key metrics providing the total number of confirmed clinical cases reported together with any increase or decrease in viral loads detected from wastewater. The dashboard also provided provincial and district level maps and viral load analyses from various WWTPs.

